# Training community home care workers to deliver a tailored falls prevention Education and Exercise program to home care clients – TrEdEx an effectiveness-implementation study protocol

**DOI:** 10.1101/2024.07.14.24310124

**Authors:** Jacqueline Francis-Coad, Anne-Marie Hill, Leon Flicker, Christopher Etherton-Beer, Elissa Burton, Philippa Wharton, Jo Wilkinson, Richard Norman, Daniel Xu, Sharmila Vaz, Vanessa Jessup

## Abstract

Frail, older people receiving supportive care in the home are at high risk of falls and functional decline that leads to either unplanned hospital admissions or permanent residential care, making it difficult to safely remain at home. Community home care organisations are well positioned to deliver high quality falls prevention programs if staff are suitably trained. This research aims to train community home care workers to deliver a fall prevention program to home care clients and evaluate program implementation and effectiveness.

A 2-phase hybrid effectiveness-implementation, pre- post design using a realist approach will be undertaken with a home care organisation. Home care workers, comprising community therapy assistants and community support workers, employed by the organisation will be trained to deliver the program by the organisation’s allied health professional staff using a train the trainer model. A multi-media falls prevention program (education and exercises) will be tailored to the client’s falls risk profile to raise falls awareness and promote physical activity and self-management. Clients receiving home care from the organisation will be recruited. Implementation of the program will be guided by the Consolidated Framework for Implementation Research, barriers and enablers will be identified at the client, staff, and organisation levels. Program effectiveness will be determined through client engagement, program satisfaction, knowledge acquisition, attitudes and enactment of falls prevention strategies, changes in functional mobility, falls and falls injuries and a cost-consequence analysis.

If successful, home care clients may enhance their functional mobility and reduce their risk of falling, allowing them to stay safely at home. Home care organisations could positively contribute to the sustainable development of a well-trained workforce delivering evidence-based programs.

## INTRODUCTION

Falls among older people remain a critical global problem to address, with national age adjusted fall injury rates in older people in Australia rising each year since 2006. ^1^ Older, frail people with complex care needs receiving support services in the home are at particularly high risk of falls injuries, further functional decline and admission to hospital or residential aged care. ^2, 3^ Over 258,000 people were receiving support services at home (HCP) in 2022-23 at a cost of $5.61 billion. ^4^ A recent Australian Government discussion paper has called for in-home care services to do more to provide older people with innovative high quality supports to remain independent and safe at home, focusing on areas such as staff training or value adding services. ^5^ Support services such as therapy and personal care are currently delivered by non-professional home care workers with diverse levels of training. ^6^ However, these staff are ideally positioned to deliver falls prevention programs as they have established rapport and trust with their older clients through time spent delivering existing services in the home. The feasibility of community home care workers delivering a falls prevention exercise program, which is an evidence-based strategy, within existing services has been established provided appropriate training was completed, but the effect on falls rates and cost of integration requires investigation. ^6^

Falls injuries are reduced among older adults who engage in evidence-based strategies such as assessment, exercise, addressing visual problems and home modifications. ^3^ However, older people show low levels of engagement in falls prevention strategies. ^7, 8^ Our research and others consistently demonstrated that older people have low awareness about fall risk, knowledge, opportunity and motivation to engage in evidence-based falls prevention strategies. These include low self-efficacy, unresolved medical problems, reluctance to accept assistance on discharge, lack of home equipment, beliefs and perceptions about falls and, in some cases, older adults absolving responsibility for recovery. ^8, 9^ New approaches to informing and assisting older people to take up falls prevention strategies include education and inclusion in the co-design of novel programs to facilitate engagement and enactment. ^6, 7, 9, 10^

Given the existing evidence we hypothesised that a tailored, novel fall prevention program, comprising education, exercise, and support to enact strategies cost-effectively delivered by trained, trusted community therapy assistants/support workers could enable older people to maximise their functional ability and reduce their risk of falling. This will support older people to remain safely at home. Therefore, the aims of this research are:

1. to implement a falls prevention program (education and exercise) delivered by trained home care workers to older people receiving supportive care in the home
2. to evaluate program effectiveness through:
  a. changes in older peoples’ functional mobility, falls and injurious falls rates.
  b. barriers and enablers to behaviour change (program uptake) at the older person, staff and organisation levels together with program costs. This will inform adaptation of the program for sustainable delivery.

## METHODS

### Design

A 2-phase hybrid effectiveness-implementation, pre-post-design was selected to enable evaluation of both implementation and effectiveness outcomes, this will also facilitate the translation of our evidence-based program into practice. ^11^ We will undertake a realist approach to evaluation. This approach seeks to provide not just a descriptive profile of an intervention’s outcomes but also to comprehensively identify how these interventions are influenced by current conditions (contexts) in triggering (mechanisms) the observed outcomes. ^12^ This is based on the realist assumption that interventions will only work in particular conditions referred to as generative or conditional causality. We will use a realist approach to identify causal pathways and determine what program (training and education/exercise) components work for whom (staff and home care clients), in what types of settings (home and organisation) and for what purpose? This aligns with research recommendations for complex interventions that suggest by adopting an approach that allows tailoring to local contexts, rather than rigid standardisation, better outcomes may be achieved. ^12^

**Figure 1.**
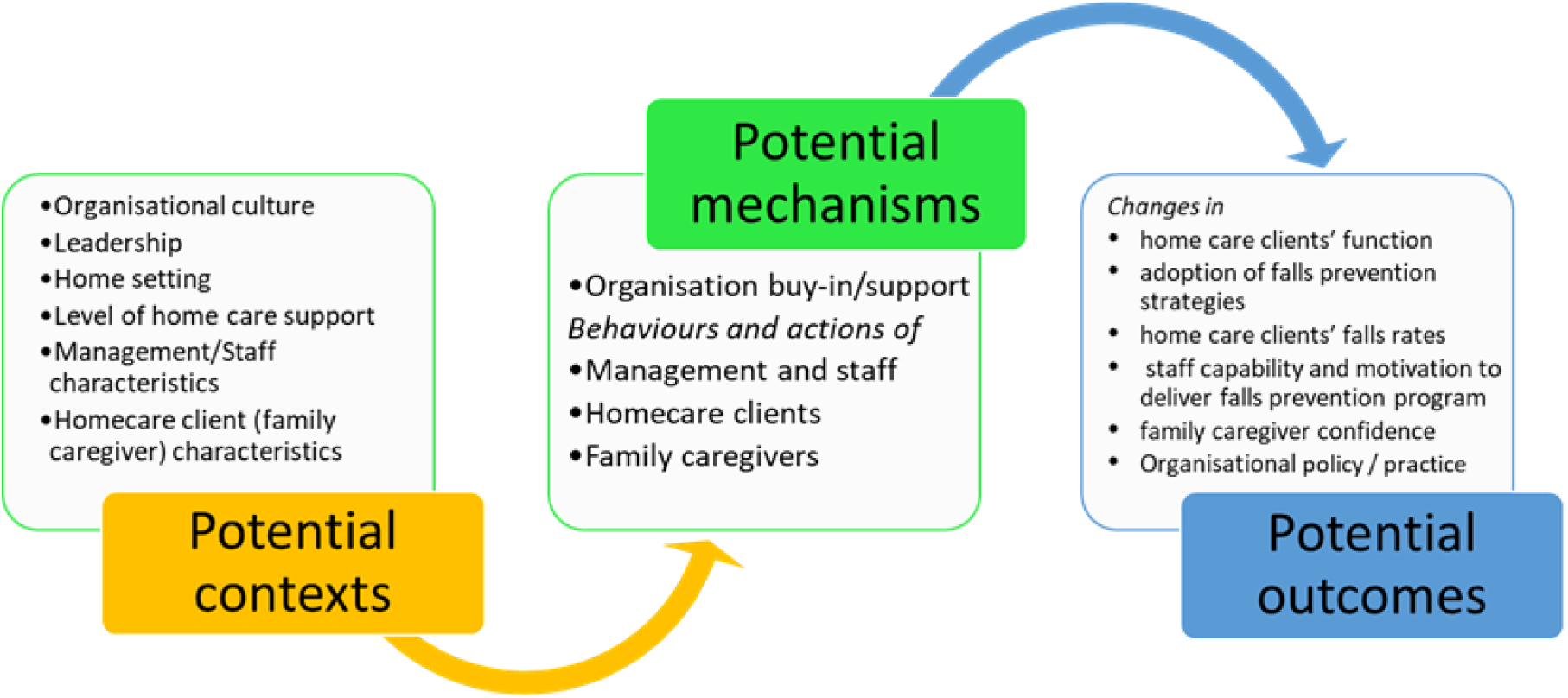
Potential context-mechanism-outcome associations

### Participants and setting

This project will partner with a large not-for-profit aged care organisation in Western Australia. The organisation provides over 575 community home care packages to older people across the Perth metropolitan area. Home care packages can deliver low to high level (1-4 respectively) support services such as assistance with showering, meals and therapy. Clients undergo the organisation’s suite of assessments on entry that include physical, cognitive and social measures. Home care managers will determine client eligibility from their organisation’s records. Eligibility criteria for home care clients will be aged 65 years and over, ambulant, receiving a home care package level 1-4 with the ability to view resources and communicate in English. Home care clients who are under a guardianship order and unable to provide consent will be excluded. If a participating client lives with a family caregiver, they will be invited to support the person they care for in undertaking the program. Home care managers will provide eligible home care clients (and their family caregiver if participating) with information regarding the study and liaise with the research team for follow up.

### Phase one

#### Data collection and procedure

##### Program review and revision

Implementing a falls prevention program into an organisation predominantly requires the program to be suitable for local needs, together with staff and clients being able to master the content and apply it consistently. ^6, 13^ We will engage purposefully sampled stakeholder groups, including home care clients, family caregivers, community therapy assistants (CTA), community support workers (CSW), allied-health professional (AHP) staff and home care managers in focus groups to determine suitability, clarity and understanding of the program content and resources. Home care managers from the organisation will identify a purposive sample of approximately 10-12 home care clients and family caregivers from their home care client database, who will be invited by telephone to the focus groups. Potential participants will be e-mailed a participant information form. Home care directors will invite approximately 10-12 staff members representing AHP, CTA, CSW, and home care managers identified from their staff records to participate in focus groups. Potential participants will be e-mailed an information form and encouraged to contact the research team to address any queries. Those committing to participate will provide written consent on arrival at the focus group. The research team (JFC, VJ) will conduct the focus groups in a private room at a convenient site using a prepared guide, conversations will be audio-recorded. Each focus group will have no more than 5-6 participants and run for approximately 1-1.5 hours. Field notes will be documented in spreadsheets on a password protected laptop computer by a research team member. Participants will also be asked to complete a short (paper) questionnaire to rate the video and printed materials. Research team members will collect all paper documents at the end of each group and place them in a sealed envelope for transportation to the university. The focus group data will provide a range of perspectives to guide tailoring of the program to meet stakeholders’ needs.

The research team will also review the organisation’s falls prevention and training policies benchmarking against evidence-based recommendations reported in the world guidelines for falls prevention and management for older adults. ^14^

##### Staff training

###### Allied health professionals

Training will form part of the organisation’s continuous quality improvement based on the organisation’s training needs analysis and commitment to providing best practice in health care. The organisation’s AHP staff (n=5), namely physiotherapists and occupational therapists, with role capacity, will be trained as program trainers. A ‘train the trainer’ model will be utilised, delivered as a 3-hour workshop by members of the research team who are experienced clinical educators (JFC, VJ & AMH). This will make delivery of the program sustainable within the organisation. These AHP staff will then be equipped to train their organisation’s CTA and CSW staff. Two of the participating AHP staff nominated by the organisation will also be trained by the research team as ‘program champions’ to co-lead the project on site, motivate staff in delivering the program, support data collection and monitor program fidelity. The train the trainer model will include:

- Adult learning principles
- Preparation for training delivery
- Facilitating training
- Problem-solving
- Role-play and simulation
- Coaching and motivation skills
- Evaluation and review

###### Home care workers

Staff comprising CTA and CSW, will deliver the falls prevention program to home care clients in phase two. The CTA staff will receive a 3-hour training workshop (face to face) delivered by the research team supported by the trained AHP staff, enabling them to deliver the falls prevention program (education and exercise) to home care clients. The CSW staff will receive a 4-hour training workshop to expand their skill set to deliver and monitor the prescribed client exercise program safely. The training will include:

- Evidence-based falls risk factor awareness and prevention management
- Coaching and motivation skills
- Goal setting and action planning
- Outcome measurement and exercise delivery

All staff nominated for training by the organisation will be invited to evaluate the training. Staff will be emailed an invitation and information form prior to the training workshops. Those volunteering to participate will sign a (paper) consent form upon on arrival at the training workshop and complete pre- and post-learning evaluation questionnaires.

### Phase two

#### Falls prevention program (intervention)

The program’s education component is based on co-designed multi-media resources previously developed and evaluated by our team, ^9, 10, 15, 16^ comprising cards/brochure and a video that address falls epidemiology, falls risk factors, actionable prevention strategies and motivation tips. Fifteen falls risk factors have been addressed from a positive perspective framed as ‘safety messages’ to assist mobility and independence. ^10, 16^ The safety messages are presented as rhymes that describe falls prevention actions with photographic images, such as switching on a light if getting up at night. The exercise component will comprise the evidence-based Otago program which includes lower limb strength and balance retraining exercises. ^17^

#### Data collection and procedure

The first eligible 100 home care clients identified by the organisation will be invited to participate, those consenting will be enrolled into the study and measured at 3 -time points: baseline, 12 weeks and 24 weeks. Enrolment is envisaged in consecutive waves (approximately 10 clients each week for 10 weeks) allowing completion in approximately 9-12 months. Research assistants will contact home care clients to complete the pre-program survey measures using an online platform (Zoom or Skype) or telephone if preferred. The organisations’ professional physiotherapy staff will assess participating home care clients and conduct the baseline functional measures to determine their falls risk profile, appropriate safety messages and Otago exercise prescription to form their personal falls prevention program. Professional physiotherapy staff will provide the trained CTA / CSW staff with the client’s program. Trained CTA / CSW staff will deliver the program to clients in a weekly 30-minute visit (Approximately 10 minutes education and 20 minutes exercise) for a 12-week intervention period. Clients’ programs will also contain exercises to complete on their own, with the expectation they will exercise in total 3-4 times weekly. The intervention period will be followed by a 12-week maintenance period in which clients will be encouraged to continue with their program. CSW staff not directly involved with delivering the falls prevention program but providing other home care services to participating clients, and family caregivers residing with participating clients will also be provided with an educational resource (paper or electronic format) to help support and motivate the participating client in adhering to their program. Participating clients will keep a program diary to record participation, goal attainment and reflections, trained CTA / CSW staff will also keep an observation diary. Clients will receive 2 support telephone calls from a research assistant across intervention and maintenance periods and all CTA / CSW staff will receive a weekly support e-mail from the program champions during the intervention phase. Baseline functional measures will be repeated at 12-weeks and 24-weeks. The organisation’s professional physiotherapy staff will conduct the repeat physical test measures and collect program diaries, research assistants will conduct the repeat survey measures. Research assistants will conduct focus groups and online interviews with staff and managers to obtain final reflections on the program implementation and effectiveness. Identifying the barriers and enablers to implementation will allow us to prepare the program for scale-up. To facilitate program expansion, we will convene a follow-up meeting to disseminate findings for stakeholders and other home care organisations.

#### Outcomes

##### Implementation outcomes

We will identify barriers and enablers to program implementation. We will conduct direct observation of program fidelity, stakeholder focus groups, surveys, and interviews, including quantitative and qualitative feedback, to measure the implementation of the fall prevention program and its outcomes. We will use the consolidated framework for implementation research underpinned with a realist evaluation approach. ^12, 13^ The implementation framework consists of five domains the intervention characteristics, internal context (organisation characteristics), external context (community home care), participant characteristics and the implementation process, that are critical for successful implementation and will be used to plan, organise and conduct implementation of the staff training and fall prevention program. ^13^ We will include home care clients, AHP staff, CTA / CSW staff and home care managers’ perceptions of the falls prevention and training programs’ acceptability and suitability. Organisational readiness for change, climate and culture, management engagement, policies and cost along with staff knowledge, awareness, attitudes and beliefs regarding the programs will be examined. Barriers and enablers will be identified at the individual (client), home care and organisation levels, this will inform adaptation of the program for sustainable delivery.

##### Effectiveness outcomes

Outcomes measured will include changes in clients’ falls prevention knowledge, attitudes and confidence using custom questionnaires. The client questionnaires were adapted from designs previously tested in research projects involving older people that evaluated an older person’s knowledge, attitudes and confidence in taking action to prevent falls. ^8, 10, 15, 16^ Family caregiver concerns regarding the older person they support sustaining a fall will be measured using the Caregiver Concern Instrument. ^18^

Changes in client function will be measured using the validated tests described in Table 1. ^19–25^

**Table 1.** Project functional effectiveness outcome measures.

| **Functional Effectiveness Outcome Measures** | **Description** |
| --- | --- |
| **6 metre-walk test** | Measures gait speed which is associated with falls risk, ability to mobilise in the community and progression of health conditions; normative values for older people available by decade<sup>19</sup> |
| **Timed up and go test** | Measures functional mobility (walking, turning and sit to stand), normative values for older people available. Higher values correlate with poorer functional mobility, falls risk and walking aid dependence<sup>20</sup> |
| **Five times sit to stand test** | Measures functional lower extremity strength transitional movement strategy Balance and falls risk; normative values for older people available by decade<sup>21</sup> |
| **4-stage balance test** | Measures static balance in 4 different standing positions by timing, then changing base of support. Not being able to hold the tandem stance (position 3) for 10 secs is an indication of increased risk of falls<sup>22</sup> |
| **360 degree turn test** | Measures dynamic balance by turning in a complete circle (360 degrees) time to complete and number of steps to complete the turn are recorded. Times > 3.8 seconds denote a critical point after which the rate of dependence is significantly increased<sup>23</sup> |
| **Short FES-I** | Measures level of concern regarding the possibility of falling when undertaking common activities. Scored 1-28 with higher scores indicating higher level of concern<sup>24</sup> |
| **EQ-5D-5L** | Measures health-related quality of life using five dimensions of health with five response levels for each dimension presented as a 5-digit index value. Self-perceived health status is measured using a visual analogue scale (VAS) scored from 0-100, where 100 points equal the best health imaginable<sup>25</sup> |

The pre and post comparator measures will demonstrate effectiveness indicated by changes in falls rates per 1000 person days and falls injuries per 1000 person days and proportion of clients who have one or more falls (data on falls will be obtained for clients for the 12 months prior to the intervention commencement).

#### Data analysis

##### Implementation outcomes

Data from focus groups, surveys, interviews, and reflection diaries will be qualitative and quantitative. Quantitative survey data will be entered into Microsoft Excel spreadsheets (Microsoft Office 2021) and analysed using Stata statistical software (StataCorp LLC, Release 18, 2023). Descriptive statistics will be used to present the main quantitative survey results. Differences in staff knowledge, awareness, confidence and motivation behaviours, pre and post receiving the training program will be examined using a dependent t-test or non-parametric alternative Wilcoxon signed rank test. ^26^ Qualitative data will be managed using NVivo software (QSR International Pty Ltd. Version 12, 2018). Open-ended qualitative responses from surveys, focus groups and reflection diaries will be subject to content analysis. ^27^ Interviews will be transcribed, coded and thematically analysed using a reflective, iterative process to determine commonly repeated patterns of meaning or themes across transcripts. ^28^ Findings from all stakeholder groups will be synthesised to create the final report.

##### Effectiveness outcomes

Linear mixed models and mixed effects negative binomial models, with random subject effects, will be used to examine longitudinal continuous and count outcomes for clients’ health outcomes of gait speed, balance, lower limb strength, fear of falling and health-related quality of life over three timepoints (baseline, 12 weeks and 24 weeks). Model results will be summarised using marginal mean estimates and 95% confidence intervals. Models will be adjusted to evaluate the cluster effect of individual home care workers on client outcomes and covariates known to affect functional ability and falls (including living situation e.g. home alone or with other, comorbidities e.g. arthritis or neurological conditions, use of a walking aid, baseline levels of exercise). The proportion of people who become fallers during the study observation period will be compared between 12 months pre-intervention- to post-intervention using logistic regression, also adjusting for the cluster effect of individual home care workers. Falls and injurious falls rates across the group during the study compared to the 12 months prior to commencement in the study will be analysed using negative binomial regression with adjustment for the home client’s length of observation in the study, and adjustment of the natural cluster of clients to individual support worker.

##### Sample size

We will use gait speed as our primary outcome variable as it is an important measure of functional change and correlates with other functional activities. A change of 0.15 metres/second (m/s) is considered a minimal clinically significant change for a group over time as evidenced from multiple patient groups. ^19^ In a previous study of older adults who completed exercise interventions, the response to gait speed within each subject group was normally distributed with a standard deviation of 0.12 m/s. ^29^ Based on estimating the standard error from these results and using pre-post intervention design in our study, 100 participants will provide 80% power to detect a change as small as 0.034 m/s (p=0.05 [2 tailed]) over time. We hypothesise that the group of participants will demonstrate a larger effect than this, more in keeping with a clinically important effect, that this study will have ample power to demonstrate.

#### Economic Evaluation

A cost-consequence analysis of the program will be conducted. This will adopt a societal perspective. It will include health system costs associated with delivering the intervention, both in the initial training phase and in the subsequent dissemination phase, predominantly the time to deliver the service to clients. To explore the cost implications of the intervention, we will include tailored surveys to track health service utilisation, in both general practice, allied health and hospitals, and will include costs incurred by the individual and their support network (including non-health costs such as travel). All interventions will be costed using standard unit pricing. As the study is focused on implementation of an intervention already demonstrated to be effective (and hence a randomised control trial is not appropriate), we propose exploring the validity of estimating risk of hospitalisation in the broader organisation cohort using existing data, matching it to baseline risk to ensure comparability. These data will allow indicative estimation of a cost-utility analysis, using the estimated incremental benefit of the program, plus publicly available utility weights sourced through the Tufts Cost Effectiveness Analysis Registry.

#### Research rigour

Triangulation of data will be achieved by using multiple sources; stakeholder focus groups, surveys, interviews, observation and reflection diaries. Member checking will be undertaken by providing participants with their written transcripts to verify accurate representation. Confirmability will be ensured through the use of verbatim participant quotations and dependability through the provision of an audit trail enabling other researchers to follow the procedures proposed. ^28^

#### Ethics

The University research team have partnered with the participating home care organisation to ensure the research priorities align with the values of the organisation. Permission to carry out the project has been obtained from the University of Western Australia Human Research Ethics Committee (2023/ET000763) and the board of the participating organisation.

## DISCUSSION

This research aims to sustainably train community home care workers to deliver a comprehensive falls prevention program to older people receiving supportive care at home and evaluate implementation and outcome effectiveness. The rationale for this research is based on Australian government initiatives aimed at enhancing home care services to reduce expensive hospital and residential aged care admissions. ^5^ This focus is particularly relevant given the increasing prevalence of falls among older individuals, especially those receiving supportive care at home. ^1, 2^ Findings may enable home care clients to remain functionally mobile and safe at home, with home care organisations playing a pivotal role in the sustainable development of a well-trained cost-effective workforce delivering evidence-based programs.

Our broad research team has led significant falls education studies in Australian community, hospital and residential aged care settings including a range of stakeholders in program development to enhance suitability and relevance. ^8–10, 15, 16^ This body of work has co-produced tailored falls prevention education programs, incorporating a suite of multi-media resources, for different populations of older people. ^8–10, 15, 16^ Providing multi-media resources caters for a range of adult learning styles with visual (video), auditory (audio), read / write (cards / brochure) and kinaesthetic (actions) modalities addressed^30^ to maximise potential engagement. Our safety messages use rhymes and graphics, which have been shown to assist in effective processing and recall of information across the lifespan. ^10, 31^ We selected the evidence-based Otago program for the exercise component as it determines exercise prescription and offers graded strength and balance retraining exercises. The Otago program has reduced the number of falls and fall-related injuries by 35% in community-dwelling older people with the greatest benefit for those aged 80 years and above, ^17^ which is synonymous with our prospective population.

Use of a realist approach and implementation evaluation framework will add depth to understanding contextual factors influencing program outcomes, ^12, 13^ enhancing potential for program transferability to similar organisations. Inclusion of a cost-consequence analysis will inform decision-makers and contribute to the allocation of finite falls prevention resources.

## CONCLUSION

To the best of our knowledge, no prior literature evaluates the implementation and effectiveness of training home care workers in delivering a tailored falls prevention education and exercise program to community home care clients. If successful home care clients may improve their functional mobility and reduce their risk of falling, enabling them to remain safely at home. Home care organisations could positively impact community falls prevention with a suitably trained work force delivering long-term evidence-based programs.

## Data Availability

This is a study protocol so no data is available yet

## Acknowledgements

Authors sincerely thank the staff and clients at Juniper community home care.

